# An observational cohort study of bronchoalveolar lavage fluid galactomannan and *Aspergillus* culture positivity in patients requiring mechanical ventilation

**DOI:** 10.1101/2024.02.07.24302392

**Authors:** Catherine A. Gao, Nikolay S. Markov, Chiagozie Pickens, Anna Pawlowski, Mengjia Kang, James M. Walter, Benjamin D. Singer, Richard G. Wunderink, NU SCRIPT Study Investigators

**Author notes:** Address correspondence to: Catherine A. Gao, MD, Instructor of Medicine, Division of Pulmonary and Critical Care, Northwestern University Feinberg School of Medicine, Simpson Querrey 5-305, 303 E Superior Street, Chicago, Illinois 60611.

## Abstract

**Rationale:** Critically ill patients who develop invasive pulmonary aspergillosis (IPA) have high mortality rates despite antifungal therapy. Diagnosis is difficult in these patients. Bronchoalveolar lavage (BAL) fluid galactomannan (GM) is a helpful marker of infection, although the optimal cutoff for IPA is unclear. We aimed to evaluate the BAL fluid GM and fungal culture results, demographics, and outcomes among a large cohort of mechanically ventilated patients with suspected pneumonia.

**Methods:** A single-center cohort study of patients enrolled in the Successful Clinical Response in Pneumonia Therapy (SCRIPT) study from June 2018 to March 2023. Demographics, BAL results, and outcomes data were extracted from the electronic health record and compared between groups of patients who grew *Aspergillus* on a BAL fluid culture, those who had elevated BAL fluid GM levels (defined as >0.5 or >0.8) but did not grow *Aspergillus* on BAL fluid culture, and those with neither.

**Results:** Of over 1700 BAL samples from 688 patients, only 18 BAL samples grew *Aspergillus*. Patients who had a BAL sample grow *Aspergillus* (n=15) were older (median 71 vs 62 years, p=0.023), had more days intubated (29 vs 11, p=0.002), and more ICU days (34 vs 15, p=0.002) than patients whose BAL fluid culture was negative for *Aspergillus* (n=672). The BAL fluid galactomannan level was higher from samples that grew *Aspergillus* on culture than those that did not (median ODI 7.08 vs 0.11, p<0.001), though the elevation of BAL fluid GM varied across BAL samples for patients who had serial sampling. Patients who grew *Aspergillus* had a similar proportion of underlying immunocompromise compared with the patients who did not, and while no statistically significant difference in overall unfavorable outcome, had longer duration of ventilation and longer ICU stays.

**Conclusions:** In this large cohort of critically ill patients with a high number of BAL samples with GM levels, we found a relatively low rate of *Aspergillus* growth. Patients who eventually grew *Aspergillus* had inconsistently elevated BAL fluid GM, and many patients with elevated BAL fluid GM did not grow *Aspergillus*. These data suggest that the pre-test probability of invasive pulmonary aspergillosis should be considered low in a general ICU population undergoing BAL evaluation to define the etiology of pneumonia. Improved scoring systems are needed to enhance pre-test probability for diagnostic test stewardship purposes.

## Introduction

Invasive pulmonary aspergillosis (IPA) is a significant cause of morbidity and mortality, particularly in immunocompromised hosts, such as those with neutropenia and a history of organ transplantation.^1^ Critically ill patients who develop invasive aspergillosis have high mortality rates despite antifungal therapy.^2^

Diagnosis of acute fungal pneumonia in critically ill patients has been difficult, and until recent years, many definitions focused primarily on immunocompromised hosts. The European Organization for Research and Treatment of Cancer/Mycoses Study Group (EORTC/MSG) proposed invasive fungal disease definitions that were divided into 1) proven - detected by blood culture or histology and culture from a normally sterile clinical site, 2) probable - having a host factor that puts the patient at risk, clinical features of disease, and positive diagnostic tests (including indirect ones such as antigen testing), and 3) possible - having a risk factor and clinical feature but lacking mycological evidence.^3^

Given the difficulty in achieving gold standard criteria for ‘proven’ IPA, diagnostics such as measurement of galactomannan (GM) have been studied. Sampling the alveolar space with bronchoalveolar lavage (BAL) is an important diagnostic tool. In addition to fungal culture, measurement of GM, a component of *Aspergillus* cell wall detected during active infection, is helpful. Nevertheless, different BAL fluid GM optical density index (ODI) cutoffs (0.5,^4^ 0.8,^5^ or 1.0^6^) have been proposed.

Other groups have proposed different algorithms with broader radiologic criteria, including any abnormality, to examine the significance of endotracheal aspirates with positive *Aspergillus* growth, with comparison to pathology data obtained from lung biopsy or autopsy as a gold standard.^7^ This approach confirmed IPA with an area under the receiver operating curve (AUROC) of 0.76, compared with only 0.57 using EORTC/MSG criteria.

Critically ill patients with severe viral pneumonia, such as influenza or SARS-CoV-2, who develop acute IPA often lack traditional host risk factors.^8^ The reported prevalence of fungal infections ranged widely from 5-27% in patients with COVID-19.^9^ A French study of 145 critically ill patients with COVID-19 (with 475 respiratory samples) found that only 20 (14%) had traditional risk factors for invasive fungal infection, but all patients in the cohort required mechanical ventilation, 75 (54%) required extracorporeal membrane oxygenation support (ECMO), and 108 (74.5%) were alive at day 30 after intensive care unit (ICU) admission.^10^

Numerous mechanisms for IPA complicating acute viral pneumonia have been hypothesized. Severe lung damage from the primary viral infection may predispose patients to secondary fungal (and bacterial) infections. Immunomodulatory therapies, such as corticosteroids and IL-6 receptor inhibitors, used to treat patients with COVID-19 may have a role in invasive fungal infections.^11^ In addition, clinical suspicion of bacterial superinfection in patients with severe viral pneumonia results in a high frequency of antibiotic use, including broad-spectrum agents.^12,13^ Prolonged exposure to broad-spectrum antibiotics promotes susceptibility to invasive fungal infections.^14^

At our center, fungal culture and GM measured in BAL fluid are part of our typical diagnostic panel for suspected pneumonia in mechanically ventilated patients. In this study, we examined a large cohort of mechanically ventilated patients undergoing BAL to evaluate the etiology of suspected pneumonia to determine the prevalence of BAL fluid GM positivity and *Aspergillus* growth. We hypothesized that patients who grew *Aspergillus* would have more risk factors such as immunocompromised status or severe viral pneumonia compared with patients who did not, and worse outcomes.

### Methods

Study setting and participants: Patients were enrolled in the Successful Clinical Response in Pneumonia Therapy (SCRIPT) Systems Biology Center, a single-site, cohort study of mechanically ventilated patients hospitalized in the Northwestern Memorial Hospital (NMH) medical intensive care unit (MICU) who underwent BAL for suspected pneumonia.^15,16^ This study was approved by the Northwestern University Institutional Review Board with study ID STU00204868. Current analysis includes patients hospitalized from June 2018 to March 2023. Patients were followed until the end of their hospitalization. Study participants were given a new identifier and enrolled at the level of hospitalization and are hereafter referred to as ‘patient.’

All patients in our MICU who underwent a BAL for suspected pneumonia were eligible for the study. Patients’ families and legal authorized representatives were approached by our research team and consented to participate in the study, which collects residual BAL fluid obtained for routine clinical care and clinical data. The research team interviews patients or families upon study enrollment and at end of the study at hospital discharge.

Our patients are routinely sampled with BAL, either bronchoscopically or non-bronchoscopically, when clinicians suspect pneumonia.^17^ These samples are typically sent for cell count and differential, amylase, bacterial and fungal cultures, BAL fluid GM, multiplex PCR (both viral and bacterial), as well as other studies when indicated. Our physicians use these data to guide antimicrobial therapy.^18^

Data compilation: Demographics (including age, gender, race, ethnicity) and outcomes (including ICU length of stay, duration of mechanical ventilation, discharge disposition) were extracted from the electronic health record (EHR) via the Northwestern Medicine Enterprise Data Warehouse.^19^ To protect patient privacy, patients from racial groups with fewer than five participants were masked as ‘Unknown.’ Manual chart review was performed for quality control of EHR pulls. Immunocompromised state (such as HIV, organ transplant, or being on chronic immunosuppressing medication, see Supplemental Material for list of criteria) were categorized by the research intake team reviewing the chart and interviewing family members. Day by day data are aggregated for ICU days at our hospital; for patients who were transferred from an outside hospital, we do not have full day by day ICU details. Missing or unavailable data are reported as such. Since we analyzed BAL fluid results from routine clinical procedures, the sampling frequency was directed by physician decisions; hence, the missingness of follow-up samples (for example, because improving patients are sampled less frequently) are inherently informative. Patients who required lung transplantation during hospitalizations were coded as having died. An unfavorable outcome was defined as patients who died during admission, or who were discharged to hospice.

A panel of critical care physician adjudicators reviewed all patient charts and categorized patients into categories based on their enrollment BAL studies and clinical syndrome. This multi-step/multi-blinded reviewer adjudication process is described in detail separately.^20^

We examined results both at the level of each BAL and at the level of each patient admission. We examined patients and BAL samples that had BAL fluid GM ODI <0.5, BAL fluid GM ODI >0.5, BAL fluid GM ODI >0.8, and those who did or did not grow *Aspergillus* on culture. We classified voriconazole, posaconazole, isavuconazonium, and amphotericin as anti-*Aspergillus* antifungal agents. Serum galactomannan and Fungitell assays for (1-3)-β-d-glucan (BDG) were also available in many patients with a high clinical suspicion of fungal infection. We examined antibacterial antibiotic exposure both in terms of days of antibiotic administration, and in terms of spectrum breadth as summarized by the Narrow Antibiotic Treatment (NAT) Score, described in more detail in our previous work.^18,21^ A NAT of -2 indicates no antibiotics, a NAT of 0 indicates antibiotics equivalent to guideline-recommended community-acquired pneumonia therapy such as ceftriaxone plus azithromycin, and higher NAT scores indicate broader antibiotic spectrum. We examined steroid administration summarized by day; to compare between different steroid agents, we converted all steroids into hydrocortisone anti-inflammatory equivalents.

Statistical analysis: Numerical values are reported in median [quartile 1, quartile 3]. Nonparametric continuous data were compared with Mann-Whitney U (MWU) tests and multiple comparisons were evaluated with Kruskal-Wallis tests. Categorical data were compared with Chi-squared tests. A p<0.05 was the cutoff for statistical significance. Analysis and visualizations were done in Python version 3.9 with *seaborn* version 0.11.2,^22^ *matplotlib* version 3.5.1,^23^ *sklearn* version 1.0.2,^24^ *scipy* version 1.7.3,^25^ and *tableone* version 0.7.10.^26^ Full code used for analysis are shared at our code repository: https://github.com/NUSCRIPT/gao_*Aspergillus*_2024. We followed the STROBE guidelines for reporting observational studies,^27^ which is available in our Supplemental Materials.

## Results

Completed hospitalization and adjudication information were available for 688 patients enrolled in SCRIPT and all were included in analysis. 209 had SARS-CoV-2 on their enrollment BAL sample, and 208 were immunocompromised, with subgroups broken out in Table 1.

**Table 1.** Cohort demographics. To protect patient privacy, patients from a race with fewer than five individuals are aggregated into the Unknown or Not Reported category. The ‘Immunocompromised’ category is inclusive of subcategories Solid Organ Transplant, Stem Cell Transplant, Leukemia, and Chemotherapy, and were categorized by the research staff upon patient enrollment in the study, whereas neutropenia during admission was summarized separately. There were no missing data for these variables.

|  |  |
| --- | --- |
| Number | 688 |
| Age, median [Q1,Q3] | 62.0 [51.0,71.0] |
| Gender, n (%) |  |
| Female | 279 (40.6) |
| Male | 409 (59.4) |
| Ethnicity, n (%) |  |
| Hispanic or Latino | 141 (20.5) |
| Not Hispanic or Latino | 515 (74.9) |
| Unknown or Not Reported | 32 (4.7) |
| Race, n (%) |  |
| Asian | 21 (3.1) |
| Black or African American | 134 (19.5) |
| Unknown or Not Reported | 126 (18.3) |
| White | 407 (59.2) |
| Admitted with COVID-19, n (%) | 209 (30.4) |
| Admitted with Influenza, n (%) | 15 (2.2) |
| Immunocompromised, n (%) | 208 (30.2) |
| Solid Organ Transplant, n (%) | 39 (5.7) |
| Stem Cell Transplant, n (%) | 36 (5.2) |
| Leukemia, n (%) | 30 (4.4) |
| Chemotherapy, n (%) | 43 (6.2) |
| Neutropenic During Admission, n (%) | 62 (9.0) |

A total of 1737 BALs were performed, 1146 of which had GM sent, and 968 of which had fungal cultures sent. At least one GM test was sent for 528 (76.7%) patients. Patients had a median [Q1,Q3] of 2 [1,3] BALs sent during their ICU hospitalization, with 1 [0,2] GM tests sent (Supplemental Figure 1). 398 samples were obtained after antifungal therapy was already administered in the ICU.

Eighteen BALs from 15 (2.2%) patients grew *Aspergillus* species on culture. Eleven of these were reported as *Aspergillus fumigatus*, and six were reported as *Aspergillus* species, and one was reported as *Aspergillus* species, not *fumigatus*. All fifteen patients had some kind of risk factor, of either underlying immunocompromise, being neutropenic during admission, or presenting with severe viral pneumonia. Ninety-seven (14.1%) patients had at least one BAL fluid GM ODI >0.5, 68 (9.9%) patients had at least one BAL fluid GM ODI >0.8, and 53 (7.7%) patients had at least one BAL fluid GM ODI >1.0 (Figure 1A). A total of 137 BALs had GM ODI > 0.5, 97 BALs had GM ODI >0.8, and 80 BALs had a GM ODI >1.0 (Figure 1B). There were 171 instances of serum galactomannan sent in our cohort during their ICU stay; the serum galactomannan was not significantly different among patients who grew *Aspergillus* compared to those who did not, median [Q1,Q3] of 0.08 [0.05,0.20] ODI compared with 0.07 [0.05, 0.14] ODI. However, the serum Fungitell assay more divergent, with a median [Q1,Q3] of 323 [286, 375] pg/mL in patients who grew *Aspergillus* compared with 31 (our lab’s lowest value) [31,292] pg/mL in patients who did not grow *Aspergillus*. There were 129 instances of serum Fungitell assay sent in our cohort during their ICU stays.

**Figure 1.**
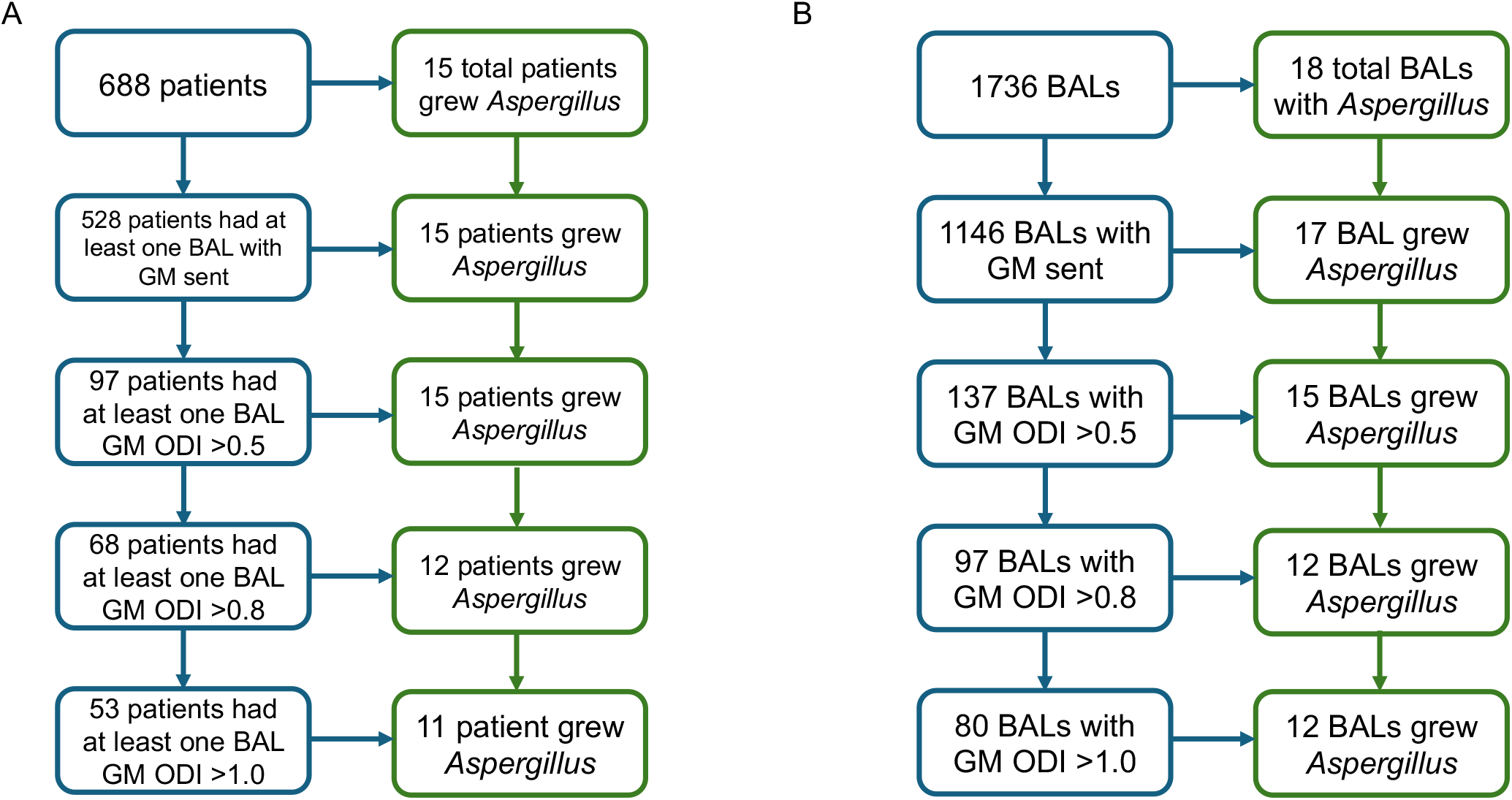
BAL fluid GM and *Aspergillus* growth. (A) Summary on a patient level. (B) Summary of BAL samples. Abbreviations: BAL: bronchoalveolar lavage, GM: galactomannan, ODI: optical density index

The median ODI of BAL fluid GM was higher in BAL samples that grew *Aspergillus* compared with those that did not: 7.08 [0.79,7.76] vs 0.11 [0.07,0.21] p<0.001. Only one patient who grew *Aspergillus* had BAL GM ODI <0.5. The 15 patients who grew *Aspergillus* had 54 BALs with GM assays sent, with median BAL fluid GM ODI of 0.78 [0.24,4.9], compared with 0.10 ([0.07,0.20], p<0.001) in the 1092 BAL samples from patients who never grew *Aspergillus* (Figure 2). Patients with BALs that grew *Aspergillus* often had elevated GM even on different BAL samples that did not grow *Aspergillus* (example timeline graphs in Figure 3).

**Figure 2.**
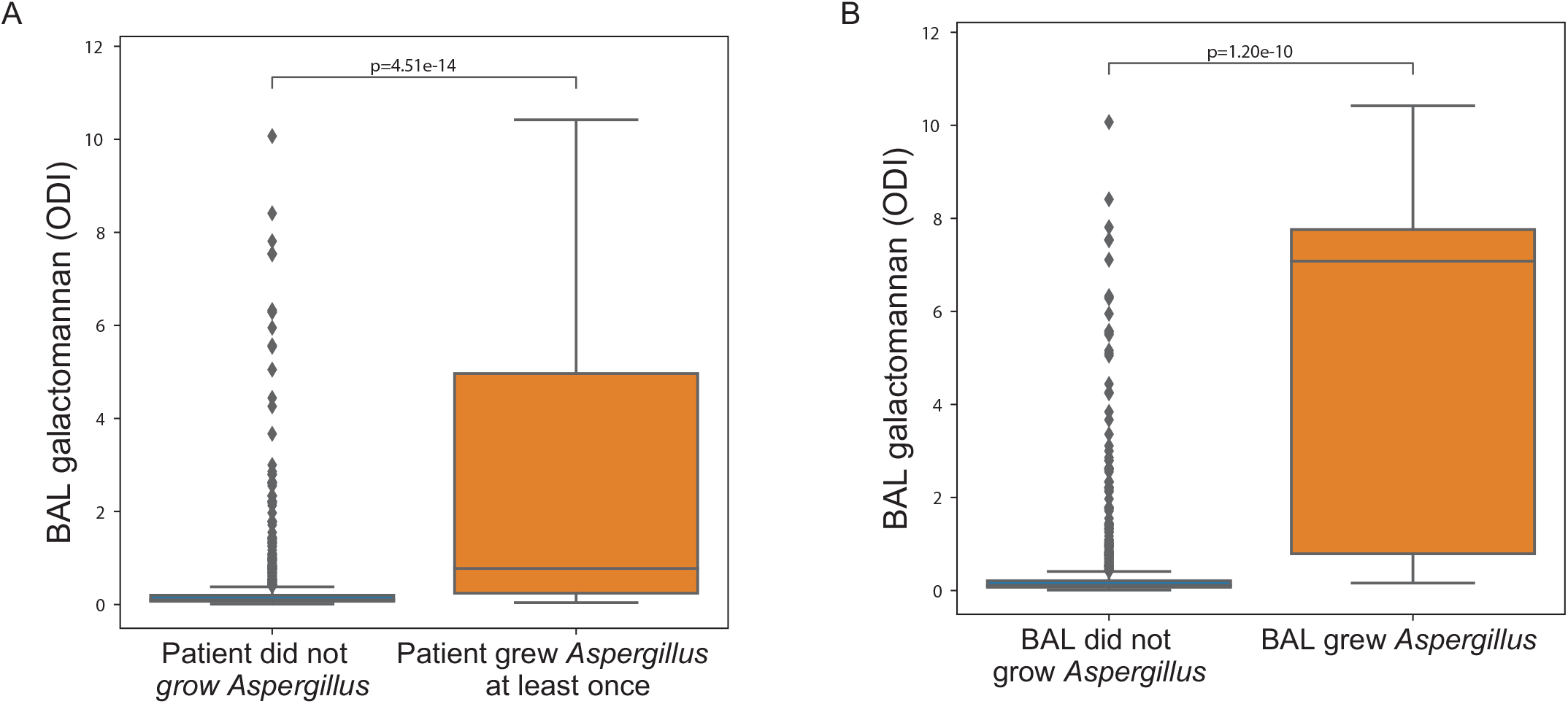
(A) BAL fluid GM ODI is higher in patients who grow *Aspergillus* at least once during admission. (B) BAL fluid GM ODI is higher in the BAL samples where *Aspergillus* grew, though sometimes also elevated in BAL samples that did not grow *Aspergillus*. Abbreviations: BAL: bronchoalveolar lavage, GM: galactomannan, ODI: optical density index

**Figure 3.**
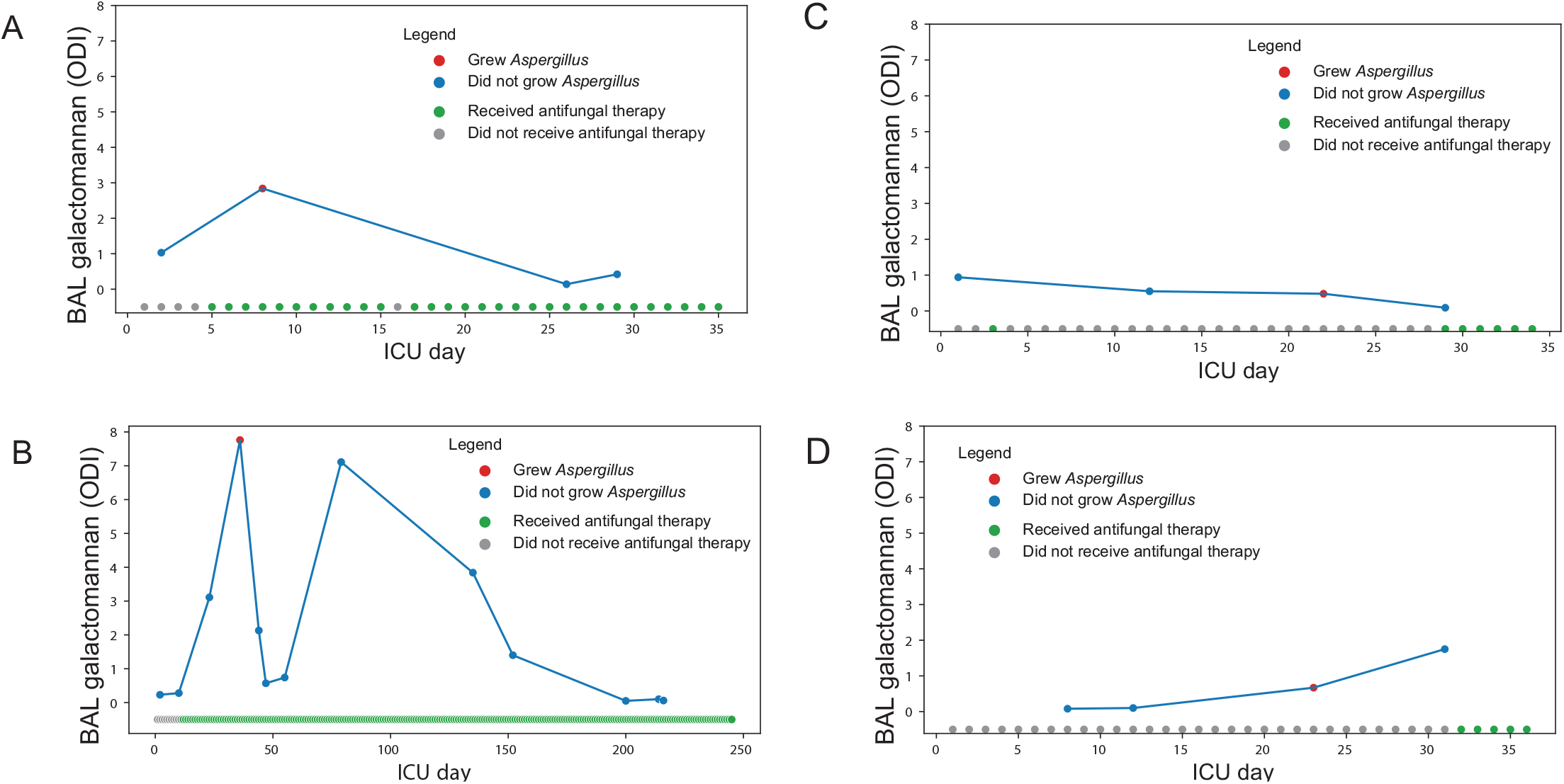
Example timelines of patients who grew *Aspergillus* on BAL fluid culture (red dot indicates *Aspergillus* growth, blue dot indicates no *Aspergillus* growth) and BAL fluid GM ODI levels (y-axis). Patients who grew *Aspergillus* did not grow it consistently, even on BAL samples that had elevated BAL fluid GM ODIs. The bottom line of each graph shows antifungal therapy by ICU day (grey = no antifungal therapy, green = antifungal therapy administered). Abbreviations: BAL: bronchoalveolar lavage, GM: galactomannan, ODI: optical density index

Eleven of the 15 patients with BAL samples that grew *Aspergillus* initially presented with COVID-19, and four were immunocompromised (two with solid organ transplant, one with chronic corticosteroid use higher than prednisone 20mg/d for the last month, and one on rituximab). Positive cultures occurred in 4.7% of patients with COVID-19 and 1.9% of immunocompromised patients in the cohort. Thirteen of the 15 patients who had BAL samples that grew *Aspergillus* were treated with antifungal therapy. One patient with a culture-positive sample who was not treated with antifungal therapy had consistently low BAL fluid GM ODI levels (0.16 and 0.05), whereas the other only had a single measurement of 0.79.

Patients with at least one BAL sample that grew *Aspergillus* (n=15) were older (71 vs 62 years, p=0.023), had more days intubated (29 vs 11 days, p=0.002) and more ICU days (34 vs 15 days, p=0.002) than patients that did not (n=672) (Table 3). They received more steroids during their ICU admission (median cumulative hydrocortisone-equivalents of 1660 vs 785mg, p=0.010). Significantly more patients with BAL that grew *Aspergillus* during their ICU stay received antifungal therapy compared to those who did not, 87% vs 37%, p<0.001. Unfavorable outcomes (death or discharge to hospice) trended higher but was not statistically different in patients with a BAL sample that grew *Aspergillus* compared to those that did not, 67% vs 45%, p=0.15.

The subset of patients with BAL fluid GM ODI >0.8 but no positive *Aspergillus* culture (n=83) was closer in age to patients without any elevated BAL fluid GM measurements, with median age of 64 years (Table 3). Their median intubation days fell between the other two categories at 17 days, as did ICU days at a median of 26 days. The percent who had a poor outcome was 52%, again between the two other categories. 64% were treated with antifungal therapy with no difference in outcome based on receipt of antifungal therapy. Results were similar for the subset of patients with elevated BAL BM ODI >0.5 (Supplemental Table 1).

*Aspergillus* first grew a median of 9 [3,22.5] days after ICU admission at our hospital, and on day 9 [2.5,22.5] of mechanical ventilation (Supplemental Table 2). The median days of antibiotics before positive culture was 8 [3,13.5] days, with cumulative NAT score of 4 [-6.5, 12.5]. Six patients had already received antifungal therapy before a positive *Aspergillus* culture. The median hydrocortisone anti-inflammatory equivalent steroid dose was 750 [520,1375] mg in the ICU days prior to growth. Among the 97 patients who had BAL fluid GM ODI>0.5 on their first sample, the sample was obtained on ICU day 2 [1,13], and ventilation day 2 [1,8]. The median days of antibiotic exposure while in the ICU before positive culture was 3 [1,9] days, with cumulative NAT score of 2 [-1,8]. Twenty-six patients had already received antifungal therapy before an elevated BAL GM. The median hydrocortisone anti-inflammatory equivalent dose was 350 [0,1300] mgs before first elevated GM.

**Table 2.** Key demographics and outcomes. Numerical values are reported as median [Q1,Q3] and compared by Mann-Whitney U tests. Categorical variables are reported as number (%) and compared by Chi-squared tests. Medication days/doses are summed across ICU days; steroid dose is in units of hydrocortisone-equivalents. Unfavorable Outcome is defined by in-hospital mortality or discharge to hospice. The ‘Immunocompromised’ category is inclusive of subcategories Solid Organ Transplant, Stem Cell Transplant, Leukemia, and Chemotherapy, and were categorized by the research staff upon patient enrollment in the study, whereas neutropenia during admission was summarized separately.

|  | Grew *Aspergillus* during admission | Did not grow *Aspergillus* | P-Value |
| --- | --- | --- | --- |
| Number | 15 | 673 |  |
| Age, median [Q1,Q3] | 71.0 [67.0,74.0] | 62.0 [51.0,71.0] | 0.023 |
| Admitted with COVID-19, n (%) | 11 (73.3) | 198 (29.4) | 0.001 |
| Admitted with Influenza, n (%) | 0 (0) | 15 (2.2) | NS |
| Immunocompromised, n (%) | 4 (26.7) | 204 (30.3) | NS |
| Solid Organ Transplant, n (%) | 2 (13.3) | 37 (5.5) | NS |
| Stem Cell Transplant, n (%) | 0 (0) | 36 (5.3) | NS |
| Leukemia, n (%) | 0 (0) | 30 (4.5) | NS |
| Chemotherapy, n (%) | 0 (0) | 43 (6.4) | NS |
| Neutropenic During Admission, n (%) | 2 (13.3) | 60 (8.9) | NS |
| Received Tocilizumab, n (%) | 1 (6.7) | 18 (2.7) | NS |
| Median PMNs over Admission, median [Q1,Q3] | 10.5 [7.2,12.9] | 8.4 [5.3,12.2] | NS |
| Cumulative ICU Days, median [Q1,Q3] | 34.0 [24.0,37.5] | 15.0 [8.0,27.0] | 0.002 |
| Cumulative Intubation Days, median [Q1,Q3] | 29.0 [23.5,36.5] | 11.0 [5.0,23.0] | 0.002 |
| Received Tracheostomy, n (%) | 8 (53.3) | 170 (25.3) | 0.031 |
| Steroid dose over ICU admission, median [Q1,Q3] | 1660.0 [1325.0,2475.0] | 780.0 [0.0,2150.0] | 0.01 |
| Treated with Antifungals, n (%) | 13 (86.7) | 249 (37.0) | <0.001 |
| Days of Antifungal Therapy, median [Q1,Q3] | 8.0 [4.5,31.0] | 0.0 [0.0,4.0] | <0.001 |
| Summed NAT Score, median [Q1,Q3] | 17.0 [-5.5,50.5] | 6.0 [-4.0,17.0] | NS |
| Days of Antibiotics Therapy, median [Q1,Q3] | 19.0 [16.0,29.0] | 11.0 [6.0,20.0] | 0.001 |
| Discharge Disposition, n (%) |  |  | NS |
| Died | 9 (60.0) | 278 (41.3) |  |
| Home | 0 (0) | 147 (21.8) |  |
| Hospice | 1 (6.7) | 23 (3.4) |  |
| LTACH | 2 (13.3) | 76 (11.3) |  |
| Rehab | 3 (20.0) | 108 (16.0) |  |
| SNF | 0 (0) | 41 (6.1) |  |
| Unfavorable Outcome, n (%) | 10 (66.7) | 301 (44.7) | NS |

**Table 3.**
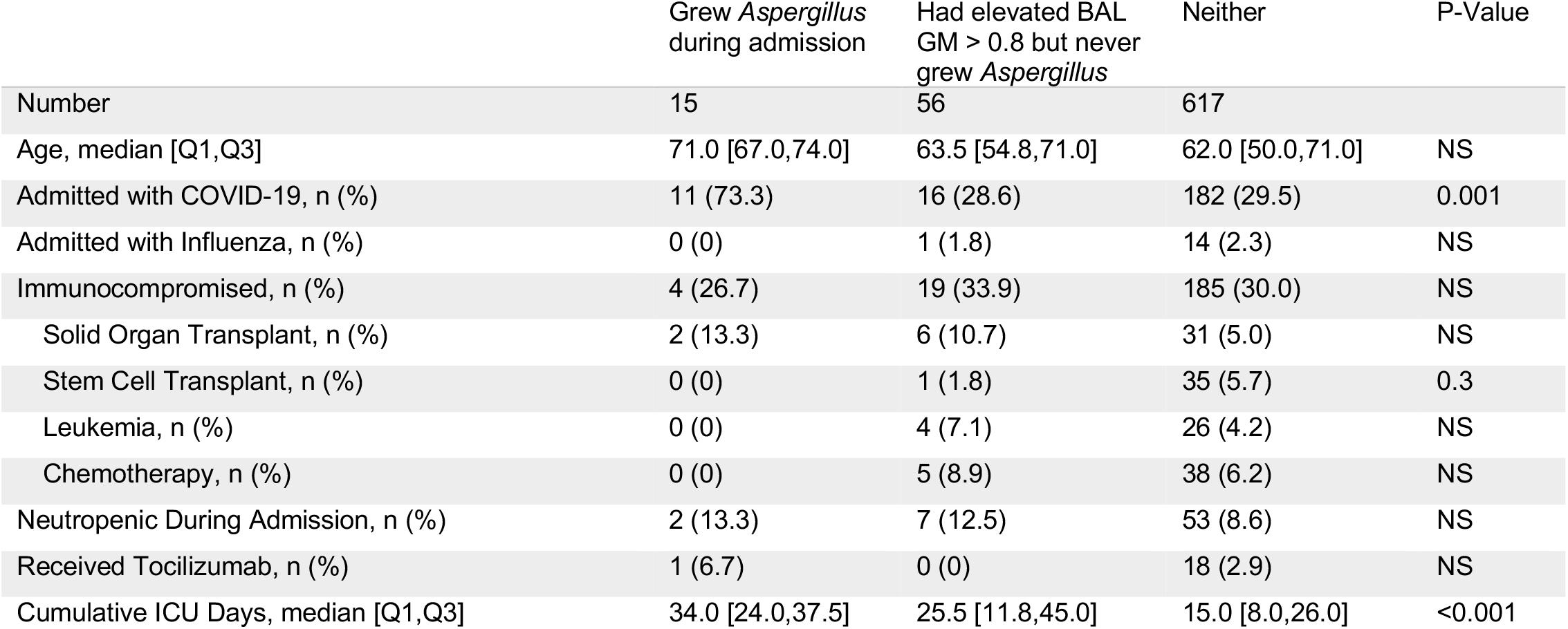
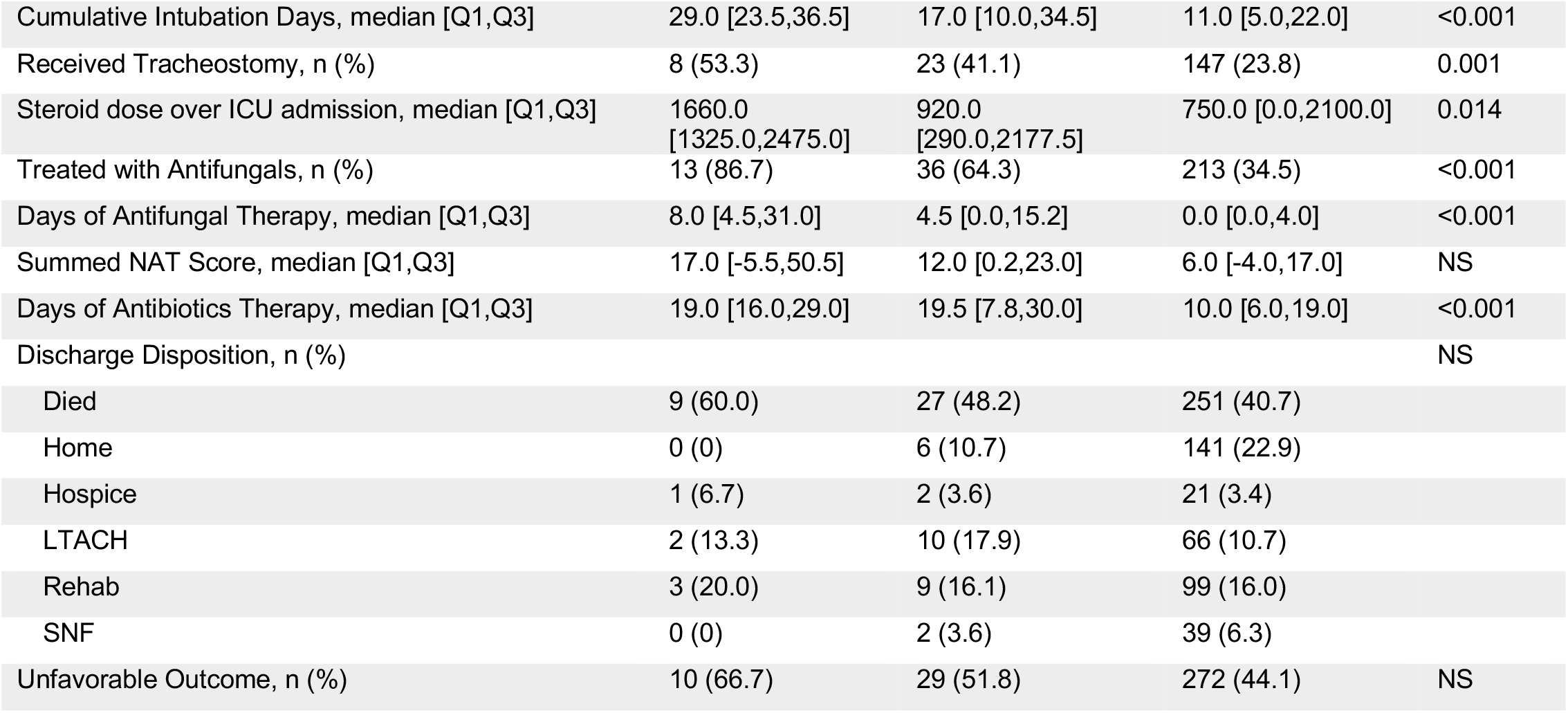
Key demographics and outcomes, split by (1) patients grew *Aspergillus* at least once during their admission, (2) patients who had elevated BAL GM > 0.8 but never grew *Aspergillus*, and (3) patients who did not have elevated BAL GM or grow *Aspergillus*. Numerical values are reported as median [Q1,Q3] and compared by Kruskal-Wallis tests. Categorical variables are reported as number (%) and compared by Chi-squared tests. Additional definitions as above in Table 2.

## Discussion

These results from a large collection of BAL samples from a cohort of mechanically ventilated patients undergoing BAL for suspected pneumonia, approximately 30% of whom were immunocompromised, demonstrated that the growth of *Aspergillus* is rare. Patients whose BAL fluid grew *Aspergillus* and patients with elevated BAL fluid GM levels, compared with patients who did not have either of these, had more ventilation days, ICU days, antibiotic days, and higher cumulative steroid dose, but no statistically significant difference in mortality. Increased mortality has been reported in other studies of *Aspergillus* superinfection, especially in studies involving patients with COVID-19-associated IPA.^28–30^ Given the small numbers of patients with *Aspergillus* growth in our cohort, our study may be underpowered to detect a statistical difference. The study cohort within SCRIPT is biased toward a more severely ill patient population compared to typical ICU patients, many of whom do not require mechanical ventilation. Even so, despite over 1700 BALs being performed, only 18 BAL samples with *Aspergillus* growth were recovered, indicating this is a rare occurrence.

Our rate of probable IPA in patients with COVID-19 is in the lower range of published rates.^31,32^ Direct comparisons are difficult across studies and case series, however, due to differences in diagnostic criteria or cutoffs, patient populations, antibiotic exposure, and environmental prevalence and exposure to *Aspergillus*. Hence, we presented our data categorized at various proposed ranges of BAL GM cutoffs. *Aspergillus* is found in a variety of environmental locations, and has various strains across the globe,^33^ possibly explaining the variability in infectious rates reported across different hospitals from different geographic locations.

A range of optimal cutoff values and algorithms have been proposed to evaluate critically ill patients for IPA.^7^ We found similar results to previous studies in that patients with positive *Aspergillus* cultures had higher BAL fluid GM compared to those who did not grow *Aspergillus*.^34^ We provide serial BAL information on many patients, which showed that patients often had elevated BAL fluid GM even on separate BALs that did not grow *Aspergillus* on culture. In contrast, elevated GM may be valuable to distinguish between colonization and active infection in samples that grow *Aspergillus* on culture. The classification of true IPA is difficult in ICU patients, as the gold standard of biopsy-proven infection often cannot be safely performed in these critically ill patients.

Most BAL fluid GM levels in this study were low, and the cohort of patients with elevated values or growth on culture was only a small proportion of the overall group. *Aspergillus* often does not grow on cultures, as shown in the multiple elevated BAL fluid GM levels in patients who only grew *Aspergillus* on one sample, or did not grow it at all. We suggest continued vigilance for *Aspergillus* superinfection in critically ill patients requiring mechanical ventilation, especially those with prolonged ventilation duration and antibiotic exposure. Patients with severe viral pneumonia such as from SARS-CoV-2 may lack traditional immunocompromised risk factors and still grow *Aspergillus*. It can be difficult to determine true IPA versus simple colonization, and the decision to treat by the clinical team was often based on judgement or patient illness. We found that serum BDG was elevated more consistently than serum GM in patients who grew *Aspergillus*, in line with previous reports of the higher sensitivity of BDG compared with GM.^35,36^

This study has several limitations. The study was conducted at a single quaternary care center with a well-established antibiotic stewardship emphasis (using rapid PCR pathogen testing to guide the narrowing antibiotics)^18^ and high use of BAL sampling compared to other centers. Given the relatively low number of patients with elevated BAL fluid GM or growth of *Aspergillus*, our study may have been underpowered to detect other important risk factors or differences in outcome. BAL GM is sent routinely by our clinical teams, irrespective of pretest probability, compared to other centers that may only send it when clinical suspicion is higher. These results and sampling practices may not reflect the population of hospitals with different antibiotic management strategies, and thus our results may not generalize to other sites. Our medication administration data was limited to doses given while in our ICU; some patients may have spent time on the hospital floor or an external ICU before being transferred to us, and medications given in those locations were not captured. Accurate radiographic studies such as CT scans to demonstrate cavitation, which suggests invasive *Aspergillus*, were not prospectively recorded in our database. However, all patients enrolled in our study had abnormal imaging that prompted a BAL for suspected pneumonia. Furthermore, it has been described that the abnormal imaging in patients with IPA without the traditional significant immunosuppression history, such as patients with COVID-19-associated pulmonary aspergillosis, lack the traditional imaging findings described with IPA (halo sign, air-crescent sign, etc.).^37^ In addition, lung abscess in ICU patients is caused by *Aspergillus* in only 8.8% of cases.^38^

In our large cohort of critically ill patients requiring mechanical ventilation, we found that *Aspergillus* growth was rare, while elevated BAL fluid GM was slightly more common and correlated with unfavorable outcomes. These data suggest that the pre-test probability of invasive pulmonary aspergillosis should be considered low in a general ICU population undergoing BAL evaluation to define the etiology of pneumonia, even among patients with immunocompromise and those with viral pneumonia. Improved scoring systems are needed to enhance pre-test probability for diagnostic test stewardship purposes.

## Supporting information

Supplemental File: STROBE checklist

Supplemental File: NU SCRIPT Study Investigators

## Data Availability

A significant portion of this data has been already made available through PhysioNet at https://physionet.org/content/script-carpediem-dataset/1.1.0/,39 a future update will include the new patients enrolled since the publication of the original dataset. Code is available at https://github.com/NUSCRIPT/gao_Aspergillus_2024.

https://physionet.org/content/script-carpediem-dataset/1.1.0/

## Data availability

A significant portion of this data has been already made available through PhysioNet at https://physionet.org/content/script-carpediem-dataset/1.1.0/,^39^ a future update will include the new patients enrolled since the publication of the original dataset. Code is available at https://github.com/NUSCRIPT/gao_Aspergillus_2024.

## Author contributions

CAG, BDS, RGW conceived and designed the study. AP and MK performed data acquisition. CAG and NSM performed data processing and cleaning. CAG performed chart review, analysis, and visualization. CAG drafted the manuscript. All authors confirm that they had full access to all the data in the study and accept responsibility to submit for publication. All authors read and approved the final draft of the manuscript.

## Supplemental Materials

Box 1. List of immunocompromised status/medications used by research team to flag patients as immunocompromised.

Medical conditions:

Acute leukemia

HIV

Immunoglobulin deficiency

Lymphoma

Multiple myeloma

Solid organ transplant

Stem cell transplant

Hematologic malign

Other malign

Non-malign

Medications:

Azathioprine

Chronic corticosteroids (last month) > 5 mg/d

Chronic corticosteroids (last month) >= 20 mg/d

Cyclosporine

Cytoxan

Mycophenolate (MMF)

Myelosuppressive chemotherapy

Rituximab

Tacrolimus

Other

## Supplemental Data

**Supplemental Figure 1.**
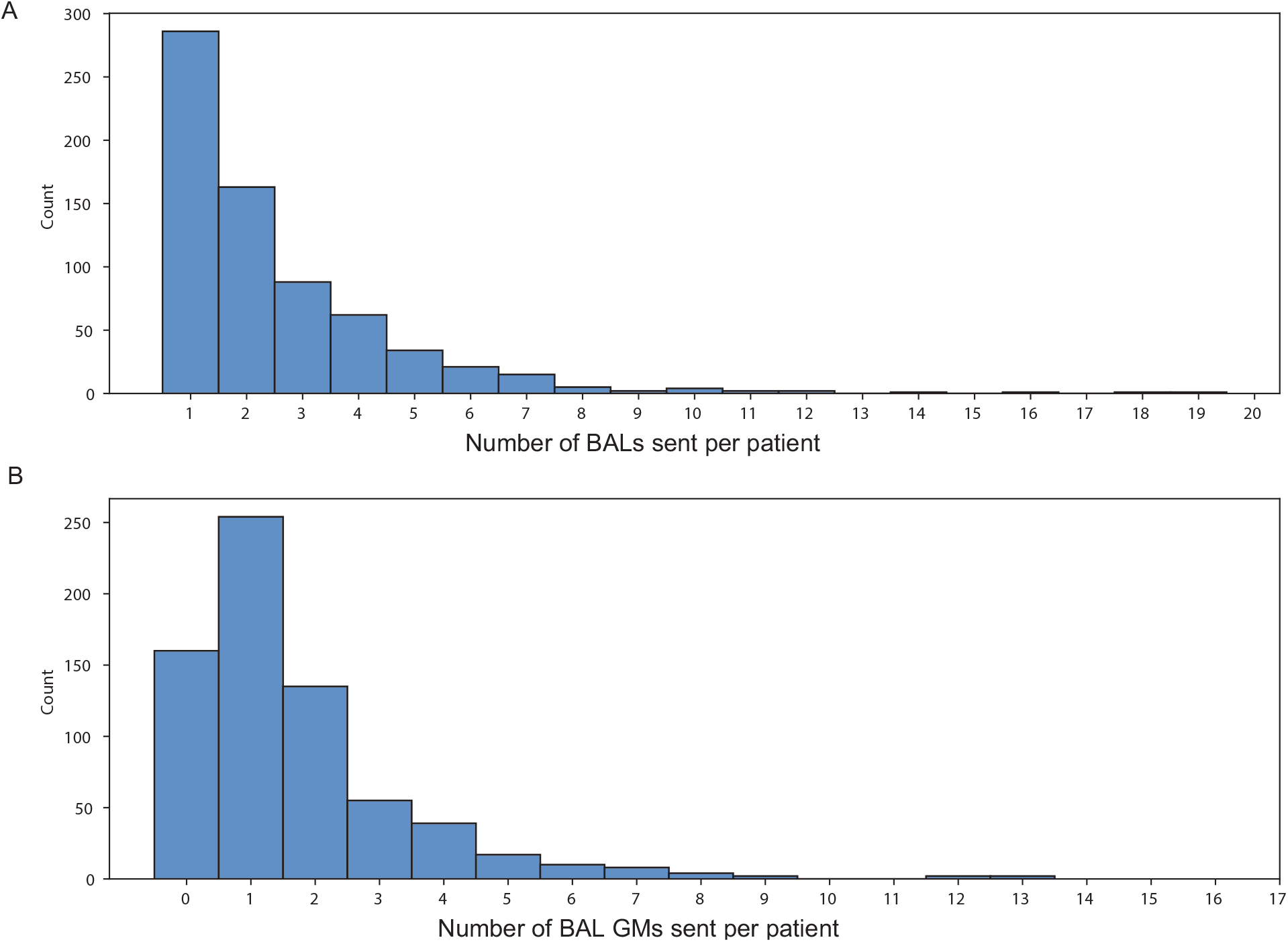
Histograms of (A) BALs performed per patient and (B) BAL fluid GMs sent per patient.

**Supplemental Table 1.**
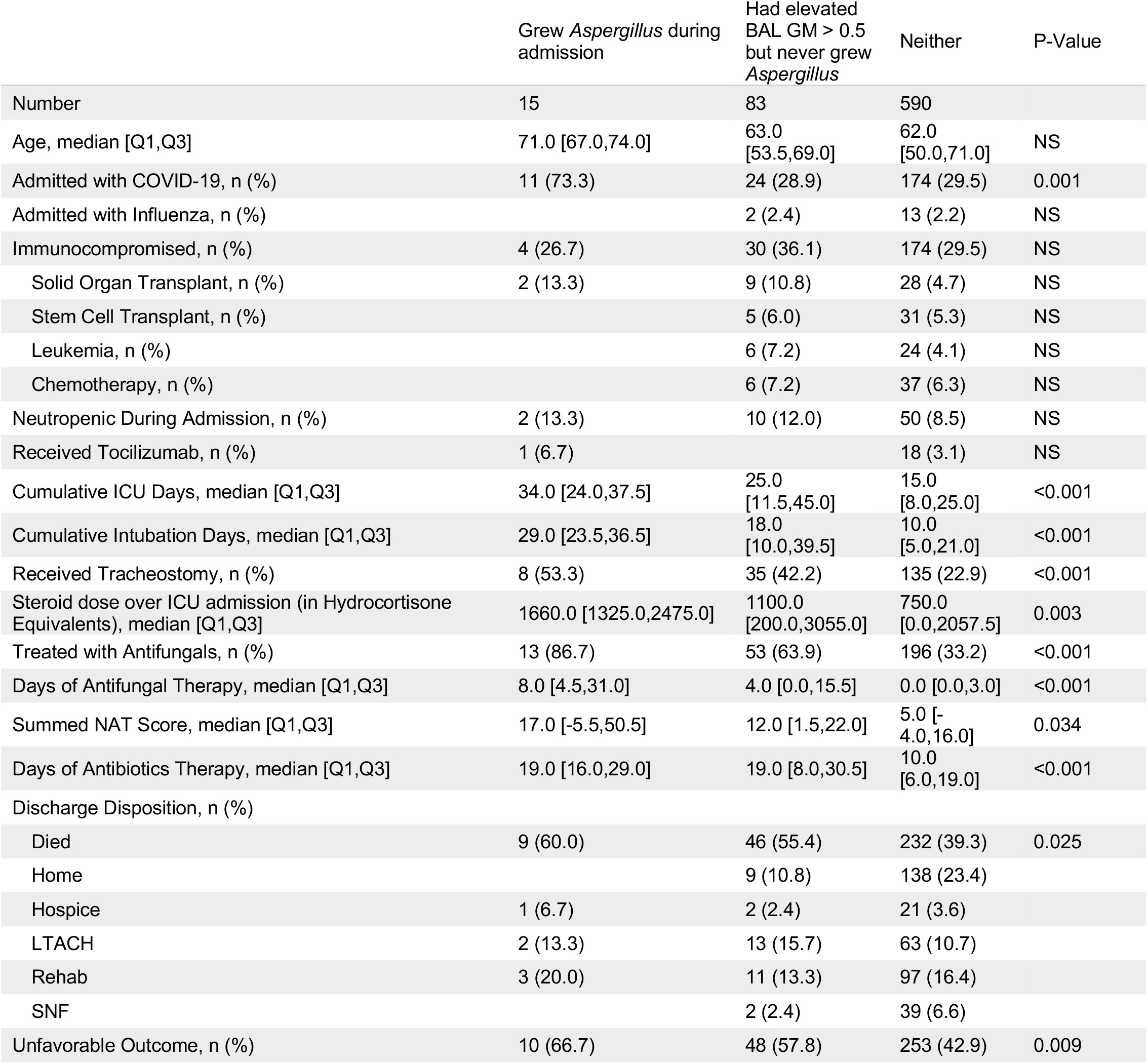
Key demographics and outcomes, split by (1) patients grew *Aspergillus* at least once during their admission, (2) patients who had elevated BAL GM > 0.5 but never grew *Aspergillus*, and (3) patients who did not have elevated BAL GM or grow *Aspergillus*. Numerical values are reported as median [Q1,Q3] and compared by Kruskal-Wallis tests. Categorical variables are reported as number (%) and compared by Chi-squared tests. Medication days/doses are summed across ICU days; steroid dose is in units of hydrocortisone-equivalents. Unfavorable Outcome is defined by in-hospital mortality or discharge to hospice. The ‘Immunocompromised’ category is inclusive of subcategories Solid Organ Transplant, Stem Cell Transplant, Leukemia, and Chemotherapy, and were categorized by the research staff upon patient enrollment in the study, whereas neutropenia during admission was summarized separately.

|  | Grew *Aspergillus* during admission | Had elevated BAL GM > 0.5 but never grew *Aspergillus* | Neither | P-Value |
| --- | --- | --- | --- | --- |
| Number | 15 | 83 | 590 |  |
| Age, median [Q1,Q3] | 71.0 [67.0,74.0] | 63.0 [53.5,69.0] | 62.0 [50.0,71.0] | NS |
| Admitted with COVID-19, n (%) | 11 (73.3) | 24 (28.9) | 174 (29.5) | 0.001 |
| Admitted with Influenza, n (%) |  | 2 (2.4) | 13 (2.2) | NS |
| Immunocompromised, n (%) | 4 (26.7) | 30 (36.1) | 174 (29.5) | NS |
| Solid Organ Transplant, n (%) | 2 (13.3) | 9 (10.8) | 28 (4.7) | NS |
| Stem Cell Transplant, n (%) |  | 5 (6.0) | 31 (5.3) | NS |
| Leukemia, n (%) |  | 6 (7.2) | 24 (4.1) | NS |
| Chemotherapy, n (%) |  | 6 (7.2) | 37 (6.3) | NS |
| Neutropenic During Admission, n (%) | 2 (13.3) | 10 (12.0) | 50 (8.5) | NS |
| Received Tocilizumab, n (%) | 1 (6.7) |  | 18 (3.1) | NS |
| Cumulative ICU Days, median [Q1,Q3] | 34.0 [24.0,37.5] | 25.0 [11.5,45.0] | 15.0 [8.0,25.0] | <0.001 |
| Cumulative Intubation Days, median [Q1,Q3] | 29.0 [23.5,36.5] | 18.0 [10.0,39.5] | 10.0 [5.0,21.0] | <0.001 |
| Received Tracheostomy, n (%) | 8 (53.3) | 35 (42.2) | 135 (22.9) | <0.001 |
| Steroid dose over ICU admission (in Hydrocortisone Equivalents), median [Q1,Q3] | 1660.0 [1325.0,2475.0] | 1100.0 [200.0,3055.0] | 750.0 [0.0,2057.5] | 0.003 |
| Treated with Antifungals, n (%) | 13 (86.7) | 53 (63.9) | 196 (33.2) | <0.001 |
| Days of Antifungal Therapy, median [Q1,Q3] | 8.0 [4.5,31.0] | 4.0 [0.0,15.5] | 0.0 [0.0,3.0] | <0.001 |
| Summed NAT Score, median [Q1,Q3] | 17.0 [-5.5,50.5] | 12.0 [1.5,22.0] | 5.0 [-4.0,16.0] | 0.034 |
| Days of Antibiotics Therapy, median [Q1,Q3] | 19.0 [16.0,29.0] | 19.0 [8.0,30.5] | 10.0 [6.0,19.0] | <0.001 |
| Discharge Disposition, n (%) |  |  |  |  |
| Died | 9 (60.0) | 46 (55.4) | 232 (39.3) | 0.025 |
| Home |  | 9 (10.8) | 138 (23.4) |  |
| Hospice | 1 (6.7) | 2 (2.4) | 21 (3.6) |  |
| LTACH | 2 (13.3) | 13 (15.7) | 63 (10.7) |  |
| Rehab | 3 (20.0) | 11 (13.3) | 97 (16.4) |  |
| SNF |  | 2 (2.4) | 39 (6.6) |  |
| Unfavorable Outcome, n (%) | 10 (66.7) | 48 (57.8) | 253 (42.9) | 0.009 |

**Supplemental Table 1.** (A) Features from a patient’s first day growing *Aspergillus*, in the 15 patients who grew it. (B) Features from a patient’s first day with an elevated BAL fluid GM ODI>0.5, in those who had it. Day_after_first_icu_day indicates what day of the ICU stay this occurred. Days_on_ventilator indicates what day of ventilation this is for the patient. Summed_days_of_abx_to_today indicates how many days of antibiotics the patient had already received in the ICU. Summed_nat_score_to_today indicates the cumulative NAT score for the patient so far during their ICU stay. Days_of_antifungal_bytoday indicates how many days of antifungal therapy the patient had already received so far during their ICU stay; this does not distinguish between prophylactic or therapeutic dosing. Sum_steroids_bytoday indicates, in hydrocortisone anti-inflammatory equivalents, what cumulative dose of steroids the patient has received.

|  | Q1 | Median | Q3 |
| --- | --- | --- | --- |
| day_after_first_icu_day | 3.0 | 9.0 | 22.5 |
| days_on_ventilator | 2.5 | 9.0 | 22.5 |
| summed_days_of_abx_to_today | 3.0 | 8.0 | 13.5 |
| summed_nat_score_to_today | -6.5 | 4.0 | 12.5 |
| days_of_antifungal_bytoday | 0.0 | 0.0 | 3.5 |
| sum_steroids_bytoday | 520.0 | 750.0 | 1375.0 |

**(B) Features from a patient's first day with elevated BAL fluid GM ODI>0.5**
|  | Q1 | Median | Q3 |
| --- | --- | --- | --- |
| day_after_first_icu_day | 1.0 | 2.0 | 13.0 |
| days_on_ventilator | 1.0 | 2.0 | 8.0 |
| summed_days_of_abx_to_today | 1.0 | 3.0 | 9.0 |
| summed_nat_score_to_today | -1.0 | 2.0 | 8.0 |
| days_of_antifungal_bytoday | 0.0 | 0.0 | 1.0 |
| sum_steroids_bytoday | 0.0 | 350.0 | 1300.0 |

